# Seasonal and Daytime Variation in Multiple Immune Parameters in Humans: Evidence from 329,261 Participants of the UK Biobank Cohort

**DOI:** 10.1101/2020.10.23.20218305

**Authors:** Cathy Wyse, G O’Malley, Andrew N. Coogan, Daniel J. Smith

## Abstract

**Background:** Seasonal disease outbreaks are perennial features of human infectious disease but the factors generating these patterns are unclear. In animal studies, seasonal and circadian (daily) rhythms in immune function generate periodicity in vulnerability to disease, although it is not known whether the same applies to humans. Making use of extensive data from the UK Biobank cohort, we investigate seasonal and daytime variability in multiple immune parameters (inflammatory markers, white blood cell counts and antibody titres), and test for associations with a wide range of environmental and lifestyle factors.

**Methods and Findings:** Markers of inflammation (CRP), and white blood cell counts were measured between 8am and 7pm over a 4-year time period in 329,261 participants in UK Biobank. Individual-level data were linked to other factors that vary over seasonal and daily cycles, including changes in day length, outdoor temperature and vitamin D at the time the blood sample was collected. Analyses were further adjusted for potentially confounding lifestyle factors. Seasonal patterns were evident in lymphocyte and neutrophil counts, and CRP, but not monocytes, and these were independent of lifestyle, demographic and environmental factors. All the immune parameters assessed demonstrated significant daytime variation that was independent of confounding factors.

**Conclusions:** At a population level, human immune parameters vary across season and across time of day, independent of multiple confounding factors. Both season and time of day are fundamental dimensions of immune function that should be considered in all studies of immuno-prophylaxis and disease transmission. Strategic alignment of human activities to seasons and times of the day when we are less susceptible to infection could be an important additional tool for limiting population-level impacts of infectious diseases.

## Introduction

Annual cycles in vulnerability to infectious disease are an established feature of human epidemiology: most respiratory viruses cause winter-time infection and polio is principally a summer-time disease.^1^ Childhood infectious diseases (meningitis, mumps, pertussis and varicella)^2^ and many of the contagious diseases that affect domestic animals^3,4^ are seasonal as are relapses in autoimmune diseases.^5,6^ The factors that mediate this seasonality are poorly understood and circannual patterns are simply an assumed component of the dynamics of infectious diseases. In addition to seasonality, animals and humans are more susceptible to infectious disease during the resting phase of their daily cycle,^7^ adding a further circadian dimension to disease vulnerability.

The axial and orbital rotations of the Earth generate predictable seasonal and daily rhythms of light and darkness. These conditions in turn generate circadian and seasonal oscillations in ambient temperature, food availability, predation and risk of infection. Evolution has equipped animals with innate timing mechanisms, or “clocks”, that synchronise physiology to these recurring periods of increased risk. The circadian clock is generated by a series of interconnected transcription–translation feedback loops that regulate the expression of a panel of clock-controlled genes.^8^ Most mammalian cells contain a molecular clock and overall rhythmicity is maintained by a master clock located in the suprachiasmatic nuclei of the hypothalamus, conferring time dependence on most physiological parameters through hormonal and neural signals.^8^ The mechanisms driving seasonality in humans are unclear, but in animals, the seasonal clock is generated by changes in thyroid hormones in the brain that respond to day length signalled by the pineal hormone melatonin.^9^

The circadian clock is entrained by the 24-hour photoperiod, while the seasonal clock entrains to day length patterns in Northern latitudes and to seasonal patterns in rain and food availability in tropical regions, where day length is constant.^10,11^ This is analogous with aspects of seasonality of the human immune system, where viral infection and immune cell numbers are associated with day length (e.g. winter peak in influenza) in Northern clines and with climatic changes in tropical regions.^6–15^

Together, the seasonal and circadian clock synchronise physiology in two dimensions of time, optimising homeostasis by anticipating changes in the environment. For example, plants^16^, fish^17^, birds^18^ and mammals^19,20^ all align their immune defence with the time of day that pathogenic and physical challenge are most likely. This conservation across the biological kingdoms is strong evidence that temporal modulation of immune function is an ancient and fundamental mechanism that has evolved to optimise survival in variable environmental conditions.

Laboratory experiments corroborate epidemiological evidence of circadian and seasonal rhythms in disease susceptibility. For example, mice are more resilient to experimental inflammatory,^21,22,23^ infectious^24–26^ and physical challenges^27^ delivered at night (their active circadian phase) or in summer.^28^ Importantly, these daily cycles in vulnerability persist in constant conditions (photoperiod, temperature, or humidity)^28,29^ and are absent in animals lacking a circadian clock,^25,30^ demonstrating clock-mediated regulation that is not driven by current environmental conditions. Similar to rodents, humans are more resistant to the effects of inflammatory or infectious challenge^31, 32^ delivered during their active circadian phase (day-time), or in summer.^33,34,35^

Circulating white blood cell counts are known to oscillate across 24h under basal conditions, reflecting distribution of cells between tissues and the periphery.^36^ Importantly, these rhythms persist in constant conditions and are absent in animals with ablated clock function^36^, indicating that they are mediated via innate circadian timing mechanisms. The extensive data collection within UK Biobank represents an unprecedented opportunity to assess seasonal and time-of-day variation in levels of human immune parameters. Here we provide new evidence of endogenous seasonal and daytime variability in human immune function at a population level and we demonstrate that these patterns are independent of a wide range of demographic, environmental and lifestyle factors.

## Methods

### Study Sample

The study sample were participants of UK Biobank, a general population cohort study that recruited over half a million UK residents continuously between 2006 and 2010, at 22 assessment centres located across the UK (UKB handbook). Eligible participants who lived within travelling distance of one of 22 UK assessment centres were identified through national health service patient registers and invited to participate by mail, resulting in a 5.5% response rate.^37^ The participants were aged between 37-73 years at the time of enrolment.

Inclusion in the present study was restricted to participants who reported having no chronic disease at the time of recruitment. Participants provided full informed consent to participate in UK Biobank. This study was covered by the generic ethical approval for UK Biobank studies from the NHS National Research Ethics Service (approval letter dated 17th June 2011, Ref 11/NW/0382) for project #26209 (PI Wyse).

### Participant Measures

Participants were invited to attend the assessment centre at a pre-booked provisional appointment time between 8am-7pm; they did not self-select the time of day of attendance but were free to reschedule if required. Baseline information was collected at the assessment centre using a questionnaire and an interview, and blood samples and physical measurements were taken. Information on the demographic status of the participants included age at baseline, sex (male/female), ethnicity (White, Black, Mixed, Chinese, Asian, Other).^38^ Participants were self-categorised as morning or evening chronotype using the question, “*Do you consider yourself to be: Definitely a morning person; More a morning than evening person; More an evening than a morning person; Definitely an evening person; or Don’t know”*. The self-reported level and duration of usual physical activity was used to derive total physical activity, measured as metabolic equivalents (MET.hours/week). A proxy of sedentary behaviour was derived from the total number of self-reported hours spent driving, using a computer and watching television each day. Smoking status was self-reported and categorized as “*never smoker*,” “*current smoker*” and “*former smoker*.” The frequency and volume of alcohol intake were self-reported. Body mass index (BMI was measured by trained UK Biobank staff using standardized methods and instruments. Habitual sleep duration was self-reported in hours per 24h.^38^

Blood samples were collected at the end of the assessment centre visit, and the time was immediately recorded on computerised system by swiping the unique barcode on the collection tube. Blood cell counts were performed within 24 hours using an LH750 haematology analyser (Coulter, Beckman Coulter, Brea, CA, USA) to determine the total number of white blood cells, plus the numbers of neutrophils, lymphocytes and monocytes, which were expressed as a percentage of the total white blood cell count. Vitamin D was measured using a chemiluminescent direct competitive immunoassay (Diasorin). Serum C reactive protein (CRP) was measured using a high-sensitivity immunoturbidimetric assay performed on a Beckman Coulter clinical chemistry analyser. A subset of participants (n=9724) were chosen at random for assessment of blood levels of antibodies against 20 infectious agents (see tables S7-8 for details of antigens). Antibody levels were measured using a Luminex high-throughput platform following validation against gold-standard assays and independent reference sera. Results of analyses were expressed as median fluoresce intensity for each antigen, and seropositivity status based on suggested thresholds. Full details of the participant measures and analysis procedures are available at www.ukbiobank.co.uk.

### Environmental Variables

Latitude and longitude were derived from the postcode of residence at 1km^2^ resolution using Open Source Geographic Information System software (QGIS Open Source Geospatial Foundation Project, http://qgis.osgeo.org). These data were combined with information on the date of attendance at the assessment center to derive the length of daylight on that day for each participant using vectorial algorithms in R-software [R version] in the ‘*insol*’ package^39^ (*insol:* Solar Radiation). Daylight was approximated over the hours (9am-7pm) of the diurnal dataset by calculating the mean zenith angle of the sun at each time and assessment centre location using the R-package “*geolight*”.^40^ The derived data were verified using information provided by the Global Monitoring Division of the US Government National Oceanic and Atmospheric Administration. Outdoor temperature was averaged for the 3 weeks preceding the date of attendance from data provided by the UK Meteorological Office for the weather station nearest to each assessment center.

### Data Analysis

Seasonal and daily variation were assessed by plotting mean values of white blood cell and CRP values against month or hour of sample collection, fitting models to describe annual and daily variation, and then investigating whether any variation was independent of confounding factors and directly related to day length.

Seasonal patterns were analyzed by fitting a linear regression model for each outcome of interest that included a sine and a cosine term of transformations of the time variable, taken as month:

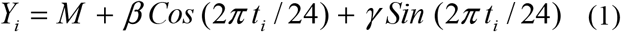

Where Y is t is time (months), and M, β and γ were predicted by regression, above. The acrophase (Φ) and amplitude (A) was predicted using equations 2 and 3, with M predicted from equation (1) above.

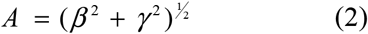

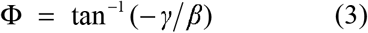

The intercept (M) was the mean level of the curve and thus an estimate of the annual mean of each outcome variable. The amplitude (A) was the distance from the mean to the acrophase or the nadir, providing an estimate of the magnitude of seasonality. The acrophase (Φ) is the peak x axis value of the curve, whereas the nadir is the trough. Seasonality was indicated by statistical significance of the estimated cosinor (sine and cosine) regression coefficients.

Variation of the markers over the daily time course of sample collection was modeled using linear methods since the absence of nighttime samples precluded assumption of circadian patterns. Although assessment centre appointments started at 8am, the blood sample was collected at the end of the 40-minute assessment, so the 8am time point was excluded due to small sample numbers at this time. The relationship between time of day and the immune parameters was represented by a series of linear regression lines connected at breakpoints where the slope of each line changed. This analysis was implemented using the R package “segmented” to predict the times of breakpoints during the test period for each analyte.^41^ The statistical significance of the segmented regression model was assessed using the Davies test to test the null hypothesis that a breakpoint does not exist, and that the difference in slope parameter (ψ) of the segmented relationship is zero. The breakpoints and slopes of each segment indicate peaks and troughs in WBC and CRP levels over time, as well as the rate and direction of any changes.

If seasonal and daily variation were indicated, we next investigated if these patterns were related to day length, and to time of day, and if any relationships were independent of lifestyle and environmental factors. The daytime data were modelled as a series of linear splines to account for the non-linear relationships between time of day and the immune parameters. Three multiple linear regression models were run that included an increasing number of covariates and progressively adjusted for sociodemographic, disease, lifestyle and environmental (temperature and day length) factors, with results reported as point estimates and 95% confidence intervals.

Potential confounders included as covariables were age; sex; ethnicity; Townsend area-deprivation score; physical activity and sedentary behaviour; alcohol intake and smoking status; outdoor temperature; blood analyser; vitamin D; sleep duration; chronotype and UK Biobank assessment centre. All analyses were performed using R version 3.5, Stata 14 statistical software (StataCorp LP) and values of p < 0.01 were considered to represent statistical significance.

## Results

The exclusion criteria for this study resulted in the removal of 173,275 study participants. The remaining cohort was mostly White (98%), with the other ethnic groups poorly represented (<2% participants). Summary data that describes the demography and lifestyle of the 329,261 participants that were eligible for inclusion are given in Table 1.

**Table 1.** Demographic and lifestyle characteristics.

|  | (n=329,261) |
| --- | --- |
| **Age (years)** |  |
| Mean (SD) | 55.8 (8.19) |
| **Townsend Deprivation Index** |  |
| Mean (SD) | -1.55 (2.94) |
| **Ethnicity** |  |
| White | 306146 (91.0%) |
| **Physical Activity** |  |
| Mean (SD) | 46.0 (62.8) |
| **Sedentary Behaviour** |  |
| Mean (SD) | 4.90 (2.24) |
| **BMI** |  |
| Mean (SD) | 26.8 (4.34) |
| **Smoking** |  |
| Yes | 31063 (9.2%) |
| **Chronotype** |  |
| More evening than morning | 81240 (24.1%) |
| Evening | 23185 (6.9%) |
| More morning than evening | 105763 (31.4%) |
| Morning | 78226 (23.2%) |

**Table 2:** Associations between day length and CRP, lymphocyte, neutrophil and monocyte count.

| Day Length | Model 1 |  | Model 2 |  | Model 3 |  |
| --- | --- | --- | --- | --- | --- | --- |
| **White Blood Cells** ($10^9$/litre) | -0.011*** | [-0.013,-0.009] | -0.008*** | [-0.011,-0.006] | -0.011*** | [-0.015,-0.007] |
| **Neutrophils** ($10^9$/litre) | -0.015*** | [-0.017,-0.014] | -0.013*** | [-0.015,-0.011] | -0.014*** | [-0.018,-0.011] |
| **Monocytes** ($10^9$/litre) | 0.001*** | [0.001,0.001] | 0.001*** | [0.001,0.001] | -0.000 | [-0.001,-0.000] |
| **Lymphocytes** ($10^9$/litre) | 0.002*** | [0.002,0.003] | 0.003*** | [0.002,0.004] | 0.004*** | [0.003,0.005] |
| **CRP** (mg/litre) | -0.006*** | [-0.007,-0.004] | -0.005*** | [-0.006,-0.003] | -0.004*** | [-0.007,-0.002] |

Data are expressed as regression coefficients (b), with 95% confidence intervals in parentheses.
Model 1 was adjusted for age, sex, ethnicity, deprivation,
Model 2 was adjusted for Model 1 + BMI, physical activity, sedentary behavior, sleep duration, chronotype, shiftwork, smoking, alcohol,
Model 3 was adjusted for Model 2 + vitamin D, outdoor temperature, time of day, blood analyser and UK Biobank assessment centre.

Mean values for WBC percentage and CRP were plotted against month and time of day for all participants, and annual and daily variation was evident on visual inspection and univariate analysis (Figures 1-2), but there was no seasonal or diurnal variation in the titre levels of any antigen (Table S7-8). The probability of seropositive status to any of the 20 antigens analysis was not associated with the month or time of day of analysis (data not shown). Summary data for white blood cell count, antigen titre, vitamin D and CRP levels at all time points are shown in Supplementary Data (Tables S1-4). Mean and 95% confidence intervals for monthly data with fitted cosinor models for lymphocyte, monocyte, neutrophil, and CRP are shown in Figure 1-2. Cosinor analysis showed that the seasonal patterns were statistically significant for a 12-month assumed periodicity for CRP, and WBC counts (Table 3).

**Table 3.** Parameters describing the amplitude, annual peak (acrophase) and mean value (mesor) predicted by fitted cosinor model. Data are mean and sd.

| Day Length | Amplitude | Acrophase | Bathyphase | Mesor | p |
| --- | --- | --- | --- | --- | --- |
| **Neutrophils** ($10^9$/litre) | 0.80 | Jan | Jul | 4.10 | <0.001 |
| **Lymphocytes** ($10^9$/litre) | 0.03 | April | Oct | 1.93 | <0.001 |
| **CRP** (mg/litre) | 0.10 | Jan | Jun | 2.23 | <0.001 |

**Figure 1:**
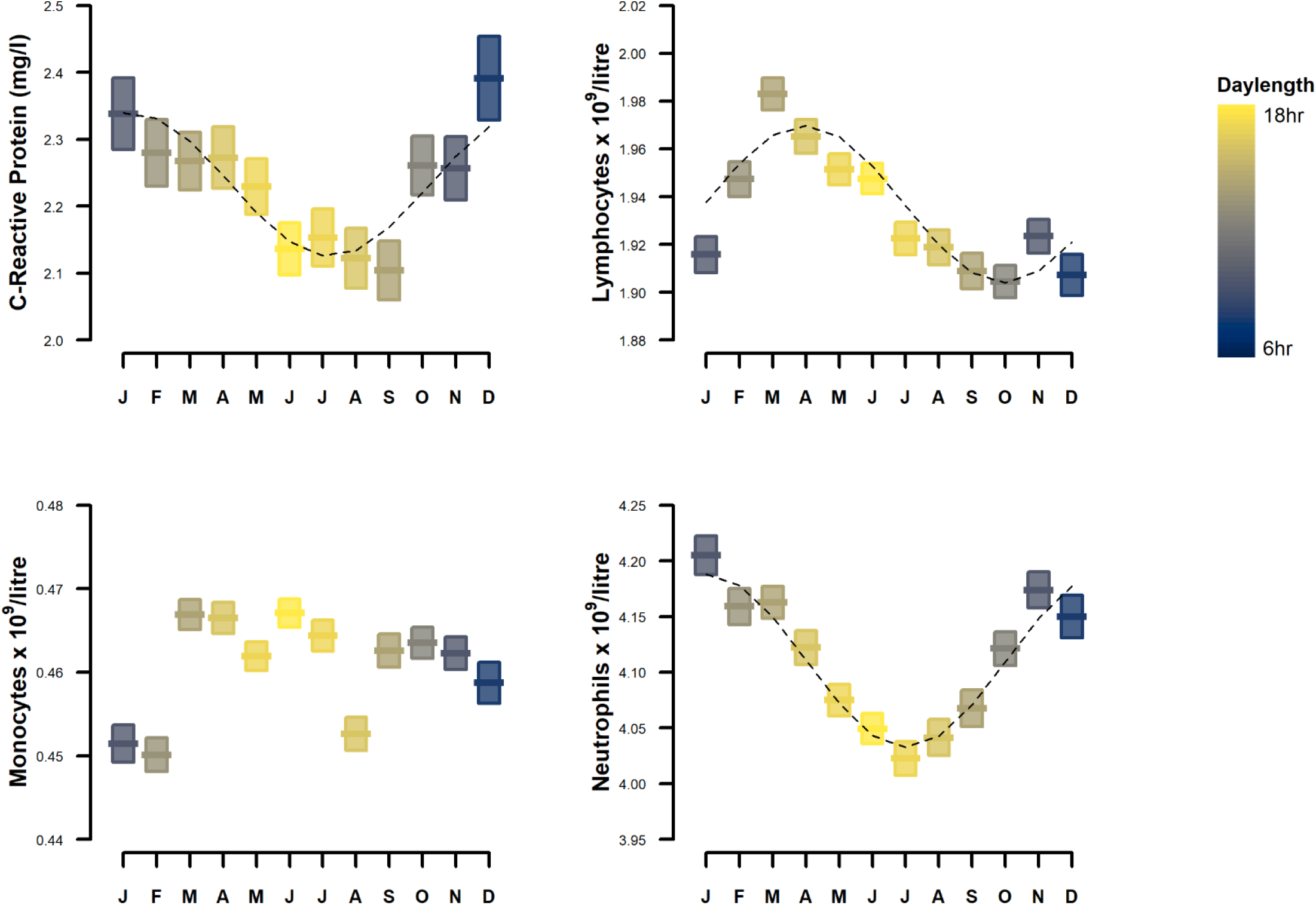
Annual variation in total monocytes, neutrophils, lymphocytes and CRP. Data are mean (bars) and 95% confidence intervals (boxes), with fitted cosinor curves (dotted line). Daylength is indicated by the box colour gradients

CRP levels were higher in the winter months, peaking in December, with lowest levels in July. The seasonal pattern of neutrophil counts was similarly higher in winter, peaking in Jan with lowest levels recorded in summer (July). The seasonal pattern of lymphocyte counts peaked in spring (March) and troughed in autumn (October) (Table 3; Figure 1). There was no significant seasonal pattern in monocyte counts.

Multiple linear regression was next applied to investigate whether the immune parameters were associated with day length and, if so, whether these were independent of other lifestyle and environmental factors that could confound associations via unrelated seasonality. CRP levels, and neutrophil and lymphocyte counts were found to be significantly associated with day length, independent of demographic, lifestyle and environmental factors (Table 2) including outdoor temperature, and vitamin D. The relationship between vitamin D and CRP was found to be dependent on BMI, and an interaction term to account for this effect was included in the CRP regression model. Interaction was also detected between vitamin D and sex for all WBC markers, and these interaction terms were added to the regression models (Supplementary Data Tables S5-6). In the fully adjusted model, neutrophil count and CRP showed significant negative associations with day length, while lymphocyte count was positively associated, as also shown in cosinor analysis. Monocyte count was not significantly associated with day length in the fully adjusted model (Model 3).

Segmented linear regression analysis of CRP and WBC counts over the daily time course showed significant daytime variation that was represented by segmental regression lines (Figure 2). The peaks and trough, (breakpoints) for each marker are shown in Table 4. All white blood cells showed significant daily variation, with counts lowest in the early morning and increasing as the day progressed. Neutrophil count reached a plateau at 3pm. CRP levels were highly variable, peaking at 1pm, and decreasing thereafter (Table 4).

**Table 4.** Segmented regression parameters showing predicted breakpoints for each segment and regression coefficient for overall segmented linear model

| Time of Day | Breakpoint<br>time of day (se) | p* |
| --- | --- | --- |
| **White Blood Cells** ($10^9$/litre) | 14.34 (0.069) | <0.001 |
| **Neutrophils** ($10^9$/litre) | 14.62 (0.040) | <0.001 |
| **Monocytes** ($10^9$/litre) | 13.27 (0.160) | <0.001 |
| **Lymphocytes** ($10^9$/litre) | 16.12 (0.180) | <0.001 |
| **CRP** (mg/litre) | 12.71 (0.220) | <0.01 |

\*Davies' test was applied to test for significant differences of slopes between each segmented relationship

**Figure 2:**
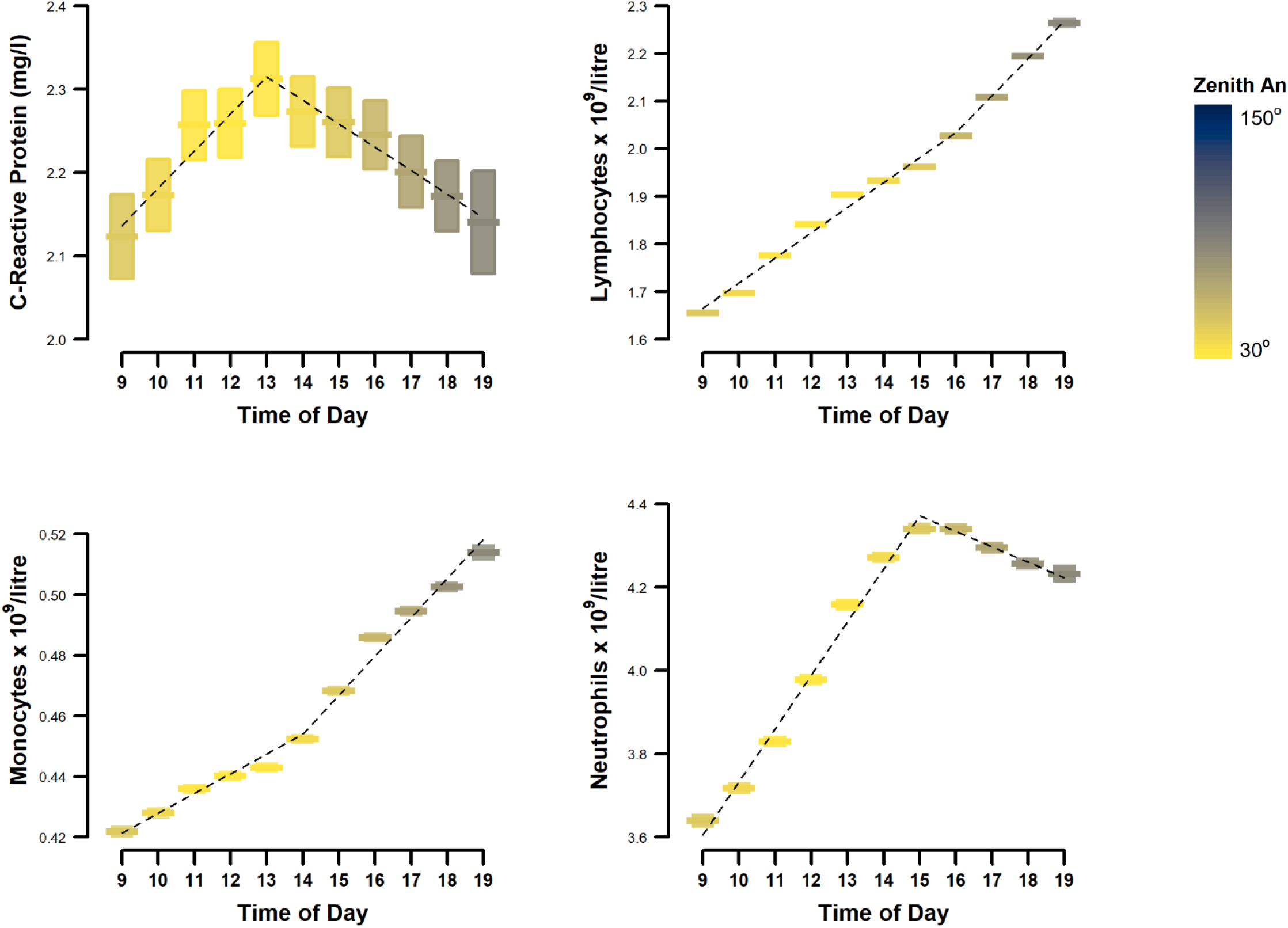
Daytime variation in monocytes, neutrophils, lymphocytes and CRP. Data are mean (bars) and 95% confidence intervals (boxes), with fitted segmented regression lines (dotted black lines). The colour gradient represents mean zenith angle of the sun at each timepoint is given to indicate daylight

Linear regression analysis demonstrated that the daytime changes in WBCs and CRP were in most cases independent of lifestyle and environmental factors (Table 5). The morning ascending segment of the CRP daily curve was the only section of any of the curves that did not retain statistical significance after adjustment. However the daily CRP curve showed a significant relationship with time of day for the later parts of the day, after the breakpoint at 1pm.

**Table 5:** Associations between time of day and CRP, lymphocyte, neutrophil and monocyte counts. Time of day is represented by 2 linear splines, to account for non-linear relationships with the independent variables.

| Day Length | Model 1 |  | Model 2 |  | Model 3 |  |
| --- | --- | --- | --- | --- | --- | --- |
| **WBCs** ($10^9$/litre) | | | | | | |
| Segment 1 | 0.207<sup>***</sup> | [0.203,0.211] | 0.199<sup>***</sup> | [0.195,0.203] | 0.197<sup>***</sup> | [0.193,0.202] |
| Segment 2 | 0.078<sup>***</sup> | [0.073,0.082] | 0.082<sup>***</sup> | [0.078,0.086] | 0.082<sup>***</sup> | [0.078,0.087] |
| **Neutrophils** ($10^9$/litre) | | | | | | |
| Segment 1 | 0.094<sup>**</sup> | [0.033,0.156] | 0.095<sup>**</sup> | [0.030,0.159] | 0.101<sup>**</sup> | [0.036,0.165] |
| Segment 2 | 0.268<sup>***</sup> | [0.243,0.293] | 0.262<sup>***</sup> | [0.236,0.289] | 0.259<sup>***</sup> | [0.233,0.285] |
| **Monocytes** ($10^9$/litre) | | | | | | |
| Segment 1 | 0.085<sup>***</sup> | [0.048,0.122] | 0.086<sup>***</sup> | [0.047,0.125] | 0.085<sup>***</sup> | [0.046,0.124] |
| Segment 2 | -0.206<sup>***</sup> | [-0.235,-0.177] | -0.199<sup>***</sup> | [-0.230,-0.168] | -0.195<sup>***</sup> | [-0.227,-0.164] |
| **Lymphocytes** ($10^9$/litre) | | | | | | |
| Segment 1 | 0.051<sup>***</sup> | [0.050,0.052] | 0.050<sup>***</sup> | [0.049,0.051] | 0.050<sup>***</sup> | [0.049,0.051] |
| Segment 2 | 0.082<sup>***</sup> | [0.079,0.085] | 0.085<sup>***</sup> | [0.082,0.088] | 0.085<sup>***</sup> | [0.082,0.088] |
| **CRP** (mg/litre) |  |  |  |  |  |  |
| Segment 1 | 0.015 | [-0.000,0.030] | 0.009 | [-0.007,0.025] | 0.010 | [-0.006,0.026] |
| Segment 2 | -0.009<sup>*</sup> | [-0.017,-0.001] | -0.013<sup>***</sup> | [-0.021,-0.006] | -0.013<sup>**</sup> | [-0.021,-0.005] |

Data are expressed as regression coefficients (b), with 95% confidence intervals in parentheses.
Model 1 was adjusted for age, sex, ethnicity, deprivation,
Model 2 was adjusted for BMI, physical activity, sedentary behaviour, sleep duration, chronotype, smoking, alcohol
Model 3 was adjusted for Model 2 + day length, blood analyser and UK Biobank assessment centre.

## Discussion

The human immune system is not constant over 24h or across the seasons and the time of exposure to pathogens is an important consideration in determining risks of infection that is relevant to public health interventions (such as movement restrictions) during epidemics.

Here we report seasonal and daytime patterns in immune cells and inflammatory markers within a large sample of the UK population. Importantly, we demonstrate that these patterns are independent of multiple demographic, lifestyle and local environmental variables, supporting the existence of endogenous seasonal and daytime patterns in human immune parameters. These findings highlight the importance of future studies to understand the time dimensions of immune function and their implications for preventing and controlling outbreaks of infectious disease.

The greatest seasonal and daytime changes in this study were seen in lymphocyte numbers, with high-amplitude variation over seasons and days. Lymphocytes were lower during the early parts of the day, increasing as the day progressed, consistent with previous reports that lymphocytes circulating in blood are lower during the respective active phase of humans^42, 43,44, 45,46^ and rodents.^43,47,48^ Circadian rhythms in the homing and egress of lymphocytes through the lymphatic system and other tissues underlies these diurnal changes of lymphocyte numbers in blood.^43,47,49^ Since the lymph nodes contain the interaction between lymphocytes and antigen, longer accumulation times increase antigen encounters and potentiate the adaptive immune response.^50,48^ Consequently, lymphocyte numbers in the periphery drop as trafficking to the tissues increases, along with increased tissue surveillance and resistance to infection.^43,49^ Circadian rhythms of lymphocyte trafficking to the periphery are abolished by genetic ablation of clock function and persist in constant conditions^47,51^ confirming their regulation by the innate circadian clock in mice. These endogenous rhythms are associated with time-of-day dependent changes in adaptive immunity, including amplified response to induction of autoimmunity (experimental autoimmune encephalomyelitis), immunisation^48^ and viral infection (influenza) during the active phase.^47^ Thus the time of day that pathogenic challenge occurs affects the adaptive immune response generated days later, one of the mechanisms through which lymphocyte trafficking might modulate seasonal and circadian vulnerability to infection. In agreement with previous findings,^52,53^ lymphocytes were positively associated with day length in our study; cell numbers were lower in autumn and peaked in spring. These findings of lower peripheral lymphocyte counts in winter and in the early active phase suggest that adaptive immune defences might be augmented in summer and during the day, and that humans might have retained some capacity for seasonal regulation of lymphocyte trafficking that might contribute to our increased susceptibility to infection in winter.

Previous studies have reported seasonal and circadian patterns in antibody titres in humans,^54^ IgM^55^, often discovered serendipitously in the course of other investigations.^55^ Leucocytes collected at different times of year showed decreased *ex vivo* response (thymidine incorporation, cytokine release) to activation in winter time in humans and rats^56–58,59^, and diurnal patterns in antibody titres^60^ and in *ex vivo* response to stimulation of PBMCs^61,62^ have been reported in humans. This study in UK Biobank is the first to investigate circadian and seasonal patterns in antibody titres to common infectious agents at a population level.

Despite our comparatively large sample size, we found no evidence for seasonal or daily variation in antibody titres or in the probability of testing immunopositive to any of the 20 antigens investigated in this study. However, the antibody response to vaccination or viral infection and subsequent decay is subject to wide variation between individuals,^63^ which is not accounted for by the cross-sectional design of the present study. Longitudinal experiments are required to establish if antibody titres vary by season or time of day and how this might impact on response to vaccination or infection. Daytime variation in antibody titres could confound studies of the efficacy of vaccination that use antibody response as an outcome variable,^60,64,65^ and future investigations within individuals and with multiple sample time points are warranted to understand basal variation in antibody titres.

Blood neutrophil counts were lowest in early morning in the UK Biobank participants, increasing thereafter to plateau after 3pm. Previous studies demonstrated comparable circadian rhythms in peripheral neutrophil counts that were low in the rest phase, and that increased over the active phase in both humans and mice.^42,66^ Neutrophils have a half-life less than 24h, and circadian rhythmicity is regulated through clock-controlled oscillations in chemokine pathways that drive release of young cells in the active phase, and clearance of aged neutrophils from the periphery in the resting phase.^67,68^ These rhythms in neutrophil tissue migration were shown to underlie increased resistance to infection (*Candidia albicans*), during the active phase in mice^68^ and to diurnal variation in bactericidal function *ex vivo* in human neutrophils.^69^ Neutrophil counts were negatively associated with day length in our study, in agreement with previous studies in humans living at temperate latitudes.^52,70^ We extend these findings to demonstrate high peripheral neutrophil counts in winter time at a population level that were related to annual photoperiod, independent of participant lifestyle, local environmental conditions and vitamin D.

In addition to total counts, previous studies have demonstrated seasonality of functional aspects of neutrophil immune function, including adhesive capacity, CD11b/CD18 expression and ROS production, resulting in augmented bactericidal properties of neutrophils collected in summer.^71^ The seasonal and daytime patterns in neutrophil count reported here, and in previous studies support evidence from animal studies that time-dependent cycles of tissue migration could contribute to neutrophil-mediated resilience to infection during the active phase, and relative vulnerability to infection in winter time.^67,68^

Monocyte counts were lower in the morning compared to evening in UK Biobank participants, consistent with previous reports that monocytes increase during the active phase in mice^72^ and humans.^45^ The acute phase protein, CRP showed a weak daily pattern in this study, with levels higher in daytime, again corroborating previous reports of diurnal patterns of CRP humans^73,52,74,75^ Circadian rhythms in circulation and tissue migration of monocytes in mice are regulated through an innate cell-intrinsic clock mechanism and their oscillation coincides with an enhanced inflammatory response when monocytes are decreasing at the beginning of the rest phase^72^ and increased lethality of endotoxic challenge at this time.^23^ In agreement, human volunteers show a heightened response to endotoxic challenge in the evening.^31,32^ This increased inflammatory response in the active phase might maximise innate immune defence at a time when pathogenic challenge is most likely,^76^ but could also leave animals vulnerable to the toxic effects of augmented inflammation.

Monocyte counts are higher in winter in some^53^ but not all^77^ previous studies. There was no evidence of a seasonal pattern in UK Biobank participants, and monocytes were not associated with day length in the fully-adjusted model. While peripheral counts are not always seasonal, monocyte function shows strong seasonality *ex vivo*, with an augmented proinflammatory response to activation in summer time.^78,79^ We found a weak seasonal pattern in the acute phase protein, CRP in UK Biobank, with levels higher in the winter months. Peripheral CRP and other proinflammatory markers were higher in winter in many studies in humans,^80,81,77^ and this is thought to contribute to seasonal prevalence of cardiovascular disease^75^

Seasonality of human viral infections is generally and intuitively thought to be driven by annual changes in temperature or humidity, but there is increasing evidence that innate variation in host disease susceptibility is an important contributor. In support of this, many diseases are seasonal in tropical regions where temperature and humidity are constant.^14,15^ Furthermore, outbreaks of influenza occur annually and simultaneously at latitudes that are oceans apart^82^ despite variations in local climatic conditions and human behaviour.

Recurrent seasonality is a feature of the epidemiology of infectious disease in animals that do not share human winter time behaviours such as increased time indoors, crowding or school terms. The prevalence of human respiratory viruses does not correspond with the prevalence of the respiratory disease they cause; remarkably, detection of viral infection is relatively low in the months that respiratory disease is highest.^83^

Vitamin D is suspected to contribute to disease seasonality due to known associations with immune function and highly seasonal serum levels,^84^ but this postulation is not corroborated by models that compared serum vitamin-D with influenza transmission in population based studies.^85,86^ Our findings in UK Biobank showed that seasonal changes in white blood cells and CRP were related to day length independent of vitamin D levels, in agreement with evidence that circulating vitamin D was not responsible for seasonality in the proinflammatory functions of human monocytes.^78^ Finally, evidence of widespread seasonal regulation of transcription of genes regulating immune function and of reversed expression patterns in Northern and Southern hemispheres strongly supports endogenous regulation of seasonality in human immune function.^87^ Seasonality of human infectious disease may be driven by an endogenous circannual rhythmicity in host immunity that generates cycles of enhancement and suppression of immune function and windows of vulnerability to infection, as proposed by Dowell (2001).^88^

This study in UK Biobank is the largest investigation of the seasonal and daytime patterns in human immune cells, inflammatory markers and antibody titres at population level, but our results are subject to many important limitations. In general, the UK Biobank does not truly represent the UK population, as participation was voluntary, uptake rates were low, and ethnic diversity was poorly represented. Furthermore, UK Biobank specifically recruited participants aged 40-69 years, so our findings may not apply to younger people. Some of the data that we analysed were self-reported, including ethnicity, physical activity, health status and chronotype, and mis-classification errors are possible. The participants denied chronic disease, but we cannot exclude the presence of acute infection at the time of assessment. The study design was cross sectional, and a single blood sample was available from each participant so the influence of within subject variation cannot be assessed. We were unable to assess circadian patterns since there were no night-time blood sample collections, and our results are limited to analysis of daytime variation. Nevertheless, the daily patterns we report in over 300,000 participants are consistent with the results of previous studies where blood was withdrawn at regular timepoints over 24h under experimental conditions. We present results of total cell counts only, and further studies are required to investigate subtypes of lymphocytes and neutrophils. The immune parameters that we report are affected by a multitude of factors related to interactions between host, pathogen and the environment. It is not possible to consider all of these in a population-based study, and the mechanisms driving the associations with day length and time of day that we report require investigation under controlled experimental conditions. Furthermore, the effect sizes we report are small, and likely to be of clinical significance for population-level disease control, rather than for the health of individuals. The strengths of this study are the large sample size and that the times of sample collection were randomly allocated to each participant. It is also a strength that we were able to investigate the effects of day length and time of day on immune parameters while adjusting for other factors thought to affect seasonal and daytime variability including physical activity, vitamin D and outdoor temperature.

Seasonality in the epidemiology of infectious disease is considered to be generated by environment and pathogen related factors, and innate variability in host susceptibility to infection is rarely considered. Our findings of seasonal and daytime variability in multiple immune parameters in a large sample of the UK population under basal, free living conditions that were independent of environmental conditions, support the contribution of innate mechanisms to variability in disease susceptibility.

Future research should focus on whether elective restriction of human activity at times of increased vulnerability to infection through night time and winter curfews could control the spread of infectious disease by minimising exposure to pathogens during susceptible periods. This is exactly the function that has driven the evolution of temporal regulation of the immune system and harnessing this innate attribute could optimise our resilience to COVID-19 and future pandemics.

## Data Availability

Data are available in the supplementary data section and at the uk biobank website

http://www.ukbiobank.co.uk

## Acknowledgements

The authors are grateful to the UK Biobank and its participants for the data used in these studies. The UK Biobank was established by the Wellcome Trust, Medical Research Council, Department of Health, Scottish Government and the Northwest Regional Development Agency. It has also had funding from the Welsh Assembly Government and the British Heart Foundation. DJS acknowledges support from a Lister Institute Prize Fellowship (2016-2021). GOM was supported by an RCSI Strategic Academic Recruitment (StAR) Fellowship. We are grateful to Claire Pallas for helpful comments on the manuscript.

## Authors Contributions

The study was conceived by CW and DS, data analysis was completed by CW. All authors contributed to writing and approved the final manuscript

## Disclosure Statement

The authors have no conflicting interests to declare.

## Data Availability

The datasets generated and/or analysed during the current study are available in the UK Biobank repository, www.ukbiobank.ac.uk. UK Biobank project: (PI: C. Wyse, Project #26209)

## Ethical Approval

The UK Biobank established a public Ethics and Governance Framework to safeguard the scientific and ethical standards of the approved research using this dataset and an independent Ethics and Governance Council to ensure that UK Biobank research adhered to the Framework. The UK Biobank project has received multiple reviews and approval from national ethical review boards in all the UK regions. In Scotland, UK Biobank has approval from the Community Health Index Advisory Group. All individuals and institutions accessing UK Biobank data are required to sign a Material Transfer Agreement that confirms that all work will be carried out in compliance with all applicable laws, regulations, ethical guidelines and approvals.

## Notes

### Competing Interest Statement

The authors have declared no competing interest.

### Funding Statement

The study received no specific funding. DS was supported by a Lister Fellowship. GOM was supported by an RCSI StAR Fellowship

### Author Declarations

UK Biobank Ethics and Governance Framework

