## supplemental for "Seasonal and Daytime Variation in Multiple Immune Parameters in Humans: Evidence from 329,261 Participants of the UK Biobank Cohort"

Table S1	Summary descriptive data for CRP, total white blood cells, monocytes, lymphocytes and neutrophils by month, mean [95% CI]

| **Month** | **January** | **February** | **March** | **April** | **May** | **June** | **July** | **August** | **September** | **October** | **November** | **December** |
| --- | --- | --- | --- | --- | --- | --- | --- | --- | --- | --- | --- | --- |
|  | *n=22269* | *n=25802* | *n=31298* | *n=27114* | *n=32005* | *n=32123* | *n=26833* | *n=23415* | *n=22680* | *n=26926* | *n=25904* | *n=17550* |
| **CRP**  mg/litre | 2.34 [2.28;2.39] | 2.28 [2.23;2.33] | 2.26 [2.22;2.31] | 2.27 [2.22;2.31] | 2.23 [2.19;2.27] | 2.13 [2.09;2.17] | 2.15 [2.11;2.19] | 2.12 [2.08;2.17] | 2.10 [2.06;2.14] | 2.26 [2.22;2.30] | 2.26 [2.21;2.30] | 2.39 [2.33;2.45] |
| **WBC**  (10^9/litre) | 6.79 [6.77;6.81] | 6.77 [6.75;6.79] | 6.83 [6.81;6.85] | 6.77 [6.75;6.79] | 6.71 [6.69;6.73] | 6.69 [6.67;6.71] | 6.63 [6.61;6.65] | 6.63 [6.61;6.65] | 6.65 [6.63;6.67] | 6.70 [6.68;6.72] | 6.78 [6.75;6.80] | 6.73 [6.70;6.75] |
| **Neutrophils** (10^9/litre) | 4.20 [4.19;4.22] | 4.16 [4.14;4.17] | 4.16 [4.15;4.17] | 4.12 [4.10;4.13] | 4.07 [4.06;4.09] | 4.05 [4.03;4.06] | 4.02 [4.01;4.04] | 4.04 [4.02;4.05] | 4.07 [4.05;4.08] | 4.12 [4.10;4.13] | 4.17 [4.16;4.19] | 4.15 [4.13;4.17] |
| **Monocytes** (10^9/litre) | 0.45 [0.45;0.45] | 0.45 [0.45;0.45] | 0.47 [0.47;0.47] | 0.47 [0.46;0.47] | 0.46 [0.46;0.46] | 0.47 [0.47;0.47] | 0.46 [0.46;0.47] | 0.45 [0.45;0.45] | 0.46 [0.46;0.46] | 0.46 [0.46;0.47] | 0.46 [0.46;0.46] | 0.46 [0.46;0.46] |
| **Lymphocytes** (10^9/litre) | 1.91 [1.91;1.92] | 1.95 [1.94;1.95] | 1.98 [1.98;1.99] | 1.97 [1.96;1.97] | 1.96 [1.95;1.96] | 1.95 [1.94;1.96] | 1.92 [1.92;1.93] | 1.92 [1.91;1.93] | 1.91 [1.90;1.92] | 1.90 [1.90;1.91] | 1.92 [1.92;1.93] | 1.91 [1.90;1.91] |

Table S2	Summary descriptive data for CRP, total white blood cells, monocytes, lymphocytes and neutrophils by time of day, mean [95% CI]

| **Time of Day** | **09.00** | **10.00** | **11.00** | **12.00** | **13.00** | **14.00** | **15.00** | **16.00** | **17.00** | **18.00** | **19.00** |
| --- | --- | --- | --- | --- | --- | --- | --- | --- | --- | --- | --- |
|  | *n=19378* | *n=27460* | *n=32984* | *n=30054* | *n=30373* | *n=32891* | *n=31529* | *n=32752* | *n=30363* | *n=30263* | *n=13013* |
| **CRP**  mg/litre | 2.12 [2.07;2.17] | 2.17 [2.13;2.22] | 2.26 [2.22;2.30] | 2.26 [2.22;2.30] | 2.31 [2.27;2.36] | 2.27 [2.23;2.31] | 2.26 [2.22;2.30] | 2.25 [2.20;2.29] | 2.20 [2.16;2.24] | 2.17 [2.13;2.21] | 2.14 [2.08;2.20] |
| **WBC**  (10^9/litre) | 5.93 [5.90;5.95] | 6.05 [6.03;6.06] | 6.24 [6.23;6.26] | 6.47 [6.45;6.48] | 6.71 [6.69;6.73] | 6.88 [6.86;6.89] | 6.99 [6.97;7.01] | 7.08 [7.06;7.10] | 7.13 [7.11;7.15] | 7.19 [7.18;7.21] | 7.25 [7.23;7.28] |
| **Neutrophils** (10^9/litre) | 3.64 [3.62;3.66] | 3.72 [3.70;3.73] | 3.83 [3.82;3.84] | 3.98 [3.96;3.99] | 4.16 [4.14;4.17] | 4.27 [4.26;4.28] | 4.34 [4.33;4.35] | 4.34 [4.33;4.35] | 4.29 [4.28;4.31] | 4.26 [4.24;4.27] | 4.23 [4.21;4.25] |
| **Monocytes** (10^9/litre) | 0.42 [0.42;0.42] | 0.43 [0.43;0.43] | 0.44 [0.43;0.44] | 0.44 [0.44;0.44] | 0.44 [0.44;0.44] | 0.45 [0.45;0.45] | 0.47 [0.47;0.47] | 0.49 [0.48;0.49] | 0.49 [0.49;0.50] | 0.50 [0.50;0.50] | 0.51 [0.51;0.52] |
| **Lymphocytes** (10^9/litre) | 1.65 [1.65;1.66] | 1.70 [1.69;1.70] | 1.78 [1.77;1.78] | 1.84 [1.84;1.85] | 1.90 [1.90;1.91] | 1.93 [1.93;1.94] | 1.96 [1.96;1.97] | 2.03 [2.02;2.03] | 2.11 [2.10;2.11] | 2.20 [2.19;2.20] | 2.26 [2.25;2.27] |

Table S3	Summary descriptive data for monocytes, lymphocytes and neutrophils expressed as percent total white blood cells by time of day, mean [95% CI]

| **Month** | **January** | **February** | **March** | **April** | **May** | **June** | **July** | **August** | **September** | **October** | **November** | **December** |
| --- | --- | --- | --- | --- | --- | --- | --- | --- | --- | --- | --- | --- |
|  | *n=22292* | *n=25833* | *n=31337* | *n=27144* | *n=32031* | *n=32162* | *n=26854* | *n=23433* | *n=22680* | *n=26952* | *n=25781* | *n=17572* |
| **Monocyte %** | 6.86 [6.82;6.89] | 6.83 [6.80;6.87] | 7.01 [6.98;7.04] | 7.06 [7.03;7.09] | 7.04 [7.02;7.07] | 7.17 [7.14;7.20] | 7.17 [7.14;7.19] | 6.99 [6.96;7.02] | 7.12 [7.09;7.15] | 7.09 [7.06;7.12] | 7.03 [7.00;7.07] | 7.01 [6.97;7.05] |
| **Lymphocyte %** | 28.8 [28.7;28.9] | 29.3 [29.2;29.4] | 29.5 [29.5;29.6] | 29.6 [29.5;29.6] | 29.7 [29.6;29.8] | 29.7 [29.6;29.8] | 29.5 [29.4;29.6] | 29.4 [29.3;29.5] | 29.2 [29.1;29.3] | 28.9 [28.9;29.0] | 29.0 [28.9;29.0] | 28.9 [28.8;29.0] |
| **Neutrophils %** | 61.3 [61.2;61.4] | 60.9 [60.8;61.0] | 60.4 [60.3;60.5] | 60.3 [60.2;60.4] | 60.2 [60.1;60.3] | 60.0 [59.9;60.1] | 60.1 [60.0;60.2] | 60.4 [60.3;60.5] | 60.6 [60.5;60.7] | 60.9 [60.8;61.0] | 61.0 [60.9;61.1] | 61.1 [60.9;61.2] |

Table S4	Summary descriptive data for monocytes, lymphocytes and neutrophils expressed as percent total white blood cells, mean [95% CI]

| **Time of Day** | **09.00** | **10.00** | **11.00** | **12.00** | **13.00** | **14.00** | **15.00** | **16.00** | **17.00** | **18.00** | **19.00** |
| --- | --- | --- | --- | --- | --- | --- | --- | --- | --- | --- | --- |
|  | *n=19374* | *n=27436* | *n=32961* | *n=30012* | *n=30350* | *n=32871* | *n=31519* | *n=32729* | *n=30335* | *n=30234* | *n=13026* |
| **Monocyte %** | 7.33 [7.29;7.37] | 7.31 [7.28;7.35] | 7.19 [7.16;7.22] | 6.98 [6.96;7.01] | 6.79 [6.76;6.82] | 6.75 [6.72;6.78] | 6.86 [6.83;6.89] | 7.02 [6.99;7.04] | 7.08 [7.05;7.10] | 7.12 [7.10;7.15] | 7.21 [7.17;7.24] |
| **Lymphocyte %** | 28.6 [28.5;28.7] | 28.7 [28.7;28.8] | 29.1 [29.0;29.2] | 29.1 [29.0;29.2] | 29.0 [28.9;29.0] | 28.7 [28.6;28.8] | 28.6 [28.5;28.7] | 29.1 [29.1;29.2] | 30.0 [29.9;30.1] | 31.0 [30.9;31.1] | 31.7 [31.5;31.8] |
| **Neutrophil %** | 60.7 [60.6;60.8] | 60.7 [60.6;60.8] | 60.6 [60.5;60.7] | 60.9 [60.8;61.0] | 61.3 [61.2;61.4] | 61.6 [61.5;61.7] | 61.6 [61.5;61.6] | 60.8 [60.7;60.9] | 59.8 [59.7;59.8] | 58.7 [58.6;58.8] | 57.8 [57.7;58.0] |

Table S5	Multiple regression analysis of associations between time of day and lymphocyte, neutrophil,

monocyte counts and CRP.

| Lymphocytes | Model 1 |  | Model 2 |  | Model 3 |  |
| --- | --- | --- | --- | --- | --- | --- |
| Segment 1 | 0.051^***^ | [0.050,0.052] | 0.050^***^ | [0.049,0.051] | 0.050^***^ | [0.049,0.051] |
| Segment 2 | 0.082^***^ | [0.079,0.085] | 0.085^***^ | [0.082,0.088] | 0.085^***^ | [0.082,0.088] |
| Sex | -0.120^***^ | [-0.124,-0.116] | -0.155^***^ | [-0.159,-0.150] | -0.154^***^ | [-0.158,-0.150] |
| Age | 0.002^***^ | [0.001,0.002] | 0.002^***^ | [0.002,0.002] | 0.002^***^ | [0.002,0.002] |
| Ethnicity | 0.047^***^ | [0.044,0.049] | 0.047^***^ | [0.044,0.051] | 0.044^***^ | [0.041,0.047] |
| Social Deprivation | 0.007^***^ | [0.006,0.007] | 0.000 | [-0.000,0.001] | -0.000 | [-0.001,0.000] |
| BMI |  |  | 0.022^***^ | [0.021,0.022] | 0.022^***^ | [0.021,0.022] |
| Physical Activity |  |  | -0.000^***^ | [-0.000,-0.000] | -0.000^***^ | [-0.000,-0.000] |
| Sedentary Behaviour |  |  | 0.009^***^ | [0.008,0.010] | 0.008^***^ | [0.007,0.009] |
| Sleep Duration |  |  | 0.006^***^ | [0.004,0.008] | 0.006^***^ | [0.004,0.008] |
| Chronotype |  |  | -0.015^***^ | [-0.017,-0.012] | -0.015^***^ | [-0.017,-0.012] |
| Smoking |  |  | 0.346^***^ | [0.339,0.353] | 0.349^***^ | [0.342,0.355] |
| Alcohol |  |  | 0.007^***^ | [0.005,0.008] | 0.007^***^ | [0.005,0.008] |
| Daylength |  |  |  |  | 0.000 | [-0.000,0.001] |
| Blood Analyser |  |  |  |  | -0.005^***^ | [-0.006,-0.003] |
| Assessment Centre |  |  |  |  | 0.004^***^ | [0.004,0.005] |
| Observations | 309220 |  | 275086 |  | 272393 |  |
| *R*^2^ | 0.099 |  | 0.159 |  | 0.161 |  |

95% confidence intervals in brackets

^*^ *p* < 0.05, ^**^ *p* < 0.01, ^***^ *p* < 0.001

| Neutrophils | Model 1 |  | Model 2 |  | Model 3 |  |
| --- | --- | --- | --- | --- | --- | --- |
| Segment1 | 0.130^***^ | [0.127,0.133] | 0.126^***^ | [0.123,0.128] | 0.125^***^ | [0.122,0.128] |
| Segment 2 | -0.039^***^ | [-0.043,-0.034] | -0.035^***^ | [-0.040,-0.030] | -0.034^***^ | [-0.039,-0.030] |
| Sex | 0.037^***^ | [0.028,0.046] | -0.021^***^ | [-0.031,-0.011] | -0.022^***^ | [-0.031,-0.012] |
| Age | -0.003^***^ | [-0.003,-0.002] | -0.002^***^ | [-0.003,-0.001] | -0.002^***^ | [-0.003,-0.001] |
| Ethnicity | -0.153^***^ | [-0.160,-0.147] | -0.146^***^ | [-0.153,-0.139] | -0.151^***^ | [-0.158,-0.144] |
| Social Deprivation | 0.021^***^ | [0.020,0.023] | 0.008^***^ | [0.007,0.010] | 0.007^***^ | [0.006,0.009] |
| BMI |  |  | 0.035^***^ | [0.034,0.036] | 0.035^***^ | [0.034,0.036] |
| Physical Activity |  |  | -0.000^***^ | [-0.000,-0.000] | -0.000^***^ | [-0.000,-0.000] |
| Sedentary Behaviour |  |  | 0.017^***^ | [0.015,0.019] | 0.016^***^ | [0.014,0.018] |
| Sleep Duration |  |  | 0.025^***^ | [0.020,0.030] | 0.025^***^ | [0.020,0.030] |
| Chronotype |  |  | -0.023^***^ | [-0.028,-0.018] | -0.023^***^ | [-0.028,-0.018] |
| Smoking |  |  | 0.794^***^ | [0.779,0.810] | 0.797^***^ | [0.781,0.813] |
| Alcohol |  |  | 0.023^***^ | [0.020,0.027] | 0.023^***^ | [0.020,0.027] |
| Daylength |  |  |  |  | -0.016^***^ | [-0.018,-0.015] |
| Blood Analyser |  |  |  |  | 0.003 | [-0.001,0.007] |
| Assessment Centre |  |  |  |  | 0.007^***^ | [0.006,0.008] |
| Observations | 308545 |  | 274483 |  | 271793 |  |
| *R*^2^ | 0.042 |  | 0.091 |  | 0.093 |  |

95% confidence intervals in brackets

^*^ *p* < 0.05, ^**^ *p* < 0.01, ^***^ *p* < 0.001

| CRP | Model 1 |  | Model 2 |  | Model 3 |  |
| --- | --- | --- | --- | --- | --- | --- |
| Segment1 | 0.013^*^ | [0.000,0.026] | 0.004 | [-0.009,0.017] | 0.005 | [-0.008,0.019] |
| Segment 2 | -0.010^*^ | [-0.019,-0.002] | -0.013^**^ | [-0.022,-0.005] | -0.013^**^ | [-0.021,-0.004] |
| Sex | -0.177^***^ | [-0.204,-0.150] | -0.402^***^ | [-0.431,-0.374] | -0.405^***^ | [-0.434,-0.377] |
| Age | 0.028^***^ | [0.026,0.029] | 0.025^***^ | [0.024,0.027] | 0.026^***^ | [0.024,0.027] |
| Ethnicity | 0.019 | [-0.002,0.039] | 0.003 | [-0.018,0.024] | 0.006 | [-0.016,0.027] |
| Social Deprivation | 0.054^***^ | [0.050,0.059] | 0.027^***^ | [0.022,0.032] | 0.029^***^ | [0.024,0.034] |
| BMI |  |  | 0.210^***^ | [0.207,0.214] | 0.210^***^ | [0.207,0.214] |
| Physical Activity |  |  | -0.001^***^ | [-0.001,-0.001] | -0.001^***^ | [-0.001,-0.001] |
| Sedentary Behaviour |  |  | 0.020^***^ | [0.014,0.026] | 0.020^***^ | [0.014,0.027] |
| Sleep Duration |  |  | 0.045^***^ | [0.031,0.059] | 0.045^***^ | [0.031,0.059] |
| Chronotype |  |  | 0.029^***^ | [0.014,0.044] | 0.029^***^ | [0.014,0.044] |
| Smoking |  |  | 0.718^***^ | [0.671,0.765] | 0.720^***^ | [0.673,0.767] |
| Alcohol |  |  | 0.014^**^ | [0.004,0.024] | 0.014^**^ | [0.004,0.024] |
| Daylength |  |  |  |  | -0.017^***^ | [-0.022,-0.013] |
| Assessment Centre |  |  |  |  | -0.005^***^ | [-0.008,-0.003] |
| Observations | 304000 |  | 270518 |  | 267872 |  |
| *R*^2^ | 0.006 |  | 0.069 |  | 0.069 |  |

95% confidence intervals in brackets

^*^ *p* < 0.05, ^**^ *p* < 0.01, ^***^ *p* < 0.001

| Monocytes | Model 1 |  | Model 2 |  | Model 3 |  |
| --- | --- | --- | --- | --- | --- | --- |
| Segment1 | 0.005^***^ | [0.005,0.006] | 0.005^***^ | [0.004,0.005] | 0.005^***^ | [0.004,0.005] |
| Segment 2 | 0.012^***^ | [0.012,0.012] | 0.012^***^ | [0.012,0.013] | 0.012^***^ | [0.012,0.013] |
| Sex | 0.070^***^ | [0.069,0.071] | 0.063^***^ | [0.062,0.064] | 0.063^***^ | [0.062,0.064] |
| Age | 0.002^***^ | [0.002,0.002] | 0.002^***^ | [0.002,0.002] | 0.002^***^ | [0.002,0.002] |
| Ethnicity | -0.015^***^ | [-0.016,-0.014] | -0.014^***^ | [-0.015,-0.014] | -0.015^***^ | [-0.016,-0.014] |
| Social Deprivation | 0.001^***^ | [0.001,0.001] | 0.000^**^ | [0.000,0.001] | 0.000 | [-0.000,0.000] |
| BMI |  |  | 0.004^***^ | [0.004,0.004] | 0.004^***^ | [0.004,0.004] |
| Physical Activity |  |  | -0.000^***^ | [-0.000,-0.000] | -0.000^***^ | [-0.000,-0.000] |
| Sedentary Behaviour |  |  | 0.001^***^ | [0.001,0.001] | 0.001^***^ | [0.001,0.001] |
| Sleep Duration |  |  | 0.002^***^ | [0.001,0.003] | 0.002^***^ | [0.002,0.003] |
| Chronotype |  |  | -0.000 | [-0.001,0.000] | -0.000 | [-0.001,0.000] |
| Smoking |  |  | 0.050^***^ | [0.048,0.052] | 0.050^***^ | [0.048,0.052] |
| Alcohol |  |  | -0.001^***^ | [-0.002,-0.001] | -0.001^***^ | [-0.002,-0.001] |
| Daylength |  |  |  |  | 0.001^***^ | [0.000,0.001] |
| Blood Analyser |  |  |  |  | -0.009^***^ | [-0.010,-0.009] |
| Assessment Centre |  |  |  |  | 0.001^***^ | [0.001,0.001] |
| Observations | 309207 |  | 275076 |  | 272380 |  |
| *R*^2^ | 0.091 |  | 0.115 |  | 0.120 |  |

95% confidence intervals in brackets

^*^ *p* < 0.05, ^**^ *p* < 0.01, ^***^ *p* < 0.001

Table S6	Multiple regression analysis of associations between daylength and lymphocyte, neutrophil,

monocyte counts and CRP.

| Neutrophils | Model 1 |  | Model 2 |  | Model 3 |  |
| --- | --- | --- | --- | --- | --- | --- |
| Daylength | 0.002^***^ | [0.002,0.003] | 0.003^***^ | [0.002,0.004] | 0.004^***^ | [0.003,0.005] |
| Sex | -0.116^***^ | [-0.120,-0.111] | -0.131^***^ | [-0.136,-0.125] | -0.129^***^ | [-0.134,-0.124] |
| Age | 0.001^***^ | [0.001,0.002] | 0.002^***^ | [0.001,0.002] | 0.001^***^ | [0.001,0.001] |
| Ethnicity | 0.056^***^ | [0.053,0.059] | 0.057^***^ | [0.053,0.060] | 0.043^***^ | [0.039,0.047] |
| Social Deprivation | 0.007^***^ | [0.006,0.008] | 0.001^*^ | [0.000,0.002] | -0.001 | [-0.002,0.000] |
| BMI |  |  | 0.022^***^ | [0.021,0.023] | 0.021^***^ | [0.020,0.022] |
| Physical Activity |  |  | -0.000^***^ | [-0.000,-0.000] | -0.000^***^ | [-0.000,-0.000] |
| Sedentary Behaviour |  |  | 0.007^***^ | [0.006,0.009] | 0.007^***^ | [0.006,0.008] |
| Sleep Duration |  |  | 0.004^**^ | [0.001,0.007] | 0.004^*^ | [0.001,0.006] |
| Chronotype |  |  | -0.007^***^ | [-0.009,-0.004] | -0.014^***^ | [-0.017,-0.011] |
| Shiftwork |  |  | -0.018^***^ | [-0.025,-0.011] | -0.009^*^ | [-0.016,-0.002] |
| Smoking |  |  | 0.334^***^ | [0.325,0.342] | 0.329^***^ | [0.321,0.338] |
| Alcohol |  |  | 0.009^***^ | [0.007,0.011] | 0.008^***^ | [0.006,0.010] |
| Vitamin D |  |  |  |  | -0.000^***^ | [-0.000,-0.000] |
| Outdoor Temperature |  |  |  |  | -0.002^***^ | [-0.003,-0.001] |
| Time of Day |  |  |  |  | 0.057^***^ | [0.057,0.058] |
| Blood Analyser |  |  |  |  | -0.005^***^ | [-0.008,-0.003] |
| Assessment Centre |  |  |  |  | 0.004^***^ | [0.004,0.005] |
| Observations | 308924 |  | 175498 |  | 161528 |  |
| *R*^2^ | 0.016 |  | 0.074 |  | 0.175 |  |

95% confidence intervals in brackets

^*^ *p* < 0.05, ^**^ *p* < 0.01, ^***^ *p* < 0.001

| Lymphocytes | Model 1 |  | Model 2 |  | Model 3 |  |
| --- | --- | --- | --- | --- | --- | --- |
| Daylength | -0.015^***^ | [-0.017,-0.014] | -0.013^***^ | [-0.015,-0.011] | -0.014^***^ | [-0.018,-0.011] |
| Sex | 0.027^***^ | [0.018,0.036] | -0.121^***^ | [-0.133,-0.108] | -0.127^***^ | [-0.140,-0.115] |
| Age | -0.000 | [-0.001,0.000] | -0.006^***^ | [-0.007,-0.005] | -0.007^***^ | [-0.008,-0.006] |
| Ethnicity | -0.141^***^ | [-0.148,-0.134] | -0.139^***^ | [-0.148,-0.131] | -0.168^***^ | [-0.178,-0.159] |
| Social Deprivation | 0.023^***^ | [0.021,0.024] | 0.005^***^ | [0.003,0.007] | 0.001 | [-0.001,0.003] |
| BMI |  |  | 0.039^***^ | [0.038,0.040] | 0.037^***^ | [0.035,0.038] |
| Physical Activity |  |  | -0.000 | [-0.000,0.000] | -0.000 | [-0.000,0.000] |
| Sedentary Behaviour |  |  | 0.018^***^ | [0.015,0.020] | 0.018^***^ | [0.015,0.021] |
| Sleep Duration |  |  | 0.023^***^ | [0.017,0.029] | 0.023^***^ | [0.017,0.030] |
| Chronotype |  |  | -0.008^*^ | [-0.015,-0.002] | -0.018^***^ | [-0.025,-0.012] |
| Shiftwork |  |  | 0.015 | [-0.001,0.032] | 0.031^***^ | [0.015,0.048] |
| Smoking |  |  | 0.773^***^ | [0.754,0.793] | 0.760^***^ | [0.740,0.780] |
| Alcohol |  |  | 0.020^***^ | [0.015,0.024] | 0.018^***^ | [0.014,0.023] |
| Vitamin D |  |  |  |  | -0.003^***^ | [-0.003,-0.002] |
| Outdoor Temperature |  |  |  |  | 0.002 | [-0.000,0.004] |
| Time of Day |  |  |  |  | 0.070^***^ | [0.068,0.072] |
| Blood Analyser |  |  |  |  | 0.000 | [-0.005,0.006] |
| Assessment Centre |  |  |  |  | 0.007^***^ | [0.006,0.008] |
| Observations | 308250 |  | 175049 |  | 161112 |  |
| *R*^2^ | 0.008 |  | 0.062 |  | 0.093 |  |

95% confidence intervals in brackets

^*^ *p* < 0.05, ^**^ *p* < 0.01, ^***^ *p* < 0.001

| Monocytes | Model 1 |  | Model 2 |  | Model 3 |  |
| --- | --- | --- | --- | --- | --- | --- |
| Daylength | 0.001^***^ | [0.001,0.001] | 0.001^***^ | [0.001,0.001] | -0.000^*^ | [-0.001,-0.000] |
| Sex | 0.071^***^ | [0.070,0.072] | 0.058^***^ | [0.056,0.059] | 0.057^***^ | [0.056,0.059] |
| Age | 0.002^***^ | [0.002,0.002] | 0.001^***^ | [0.001,0.001] | 0.001^***^ | [0.001,0.001] |
| Ethnicity | -0.013^***^ | [-0.014,-0.013] | -0.013^***^ | [-0.014,-0.012] | -0.016^***^ | [-0.017,-0.015] |
| Social Deprivation | 0.001^***^ | [0.001,0.002] | 0.000 | [-0.000,0.000] | -0.000 | [-0.000,0.000] |
| BMI |  |  | 0.004^***^ | [0.004,0.004] | 0.004^***^ | [0.004,0.004] |
| Physical Activity |  |  | -0.000 | [-0.000,0.000] | -0.000 | [-0.000,0.000] |
| Sedentary Behaviour |  |  | 0.001^***^ | [0.001,0.002] | 0.001^***^ | [0.001,0.002] |
| Sleep Duration |  |  | 0.002^***^ | [0.001,0.003] | 0.002^***^ | [0.001,0.002] |
| Chronotype |  |  | 0.001^*^ | [0.000,0.002] | -0.001 | [-0.001,0.000] |
| Shiftwork |  |  | 0.000 | [-0.002,0.002] | 0.001 | [-0.000,0.003] |
| Smoking |  |  | 0.048^***^ | [0.046,0.050] | 0.048^***^ | [0.045,0.050] |
| Alcohol |  |  | -0.001^***^ | [-0.002,-0.001] | -0.002^***^ | [-0.002,-0.001] |
| Vitamin D |  |  |  |  | -0.000^***^ | [-0.000,-0.000] |
| Outdoor Temperature |  |  |  |  | 0.001^***^ | [0.001,0.001] |
| Time of Day |  |  |  |  | 0.010^***^ | [0.009,0.010] |
| Blood Analyser |  |  |  |  | -0.010^***^ | [-0.010,-0.009] |
| Assessment Centre |  |  |  |  | 0.001^***^ | [0.001,0.001] |
| Observations | 308911 |  | 175441 |  | 161471 |  |
| *R*^2^ | 0.060 |  | 0.075 |  | 0.119 |  |

95% confidence intervals in brackets

^*^ *p* < 0.05, ^**^ *p* < 0.01, ^***^ *p* < 0.001

| CRP | Model 1 |  | Model 2 |  | Model 3 |  |
| --- | --- | --- | --- | --- | --- | --- |
| Daylength | -0.006^***^ | [-0.007,-0.004] | -0.005^***^ | [-0.006,-0.003] | -0.004^***^ | [-0.007,-0.002] |
| Sex | -0.042^***^ | [-0.050,-0.035] | -0.153^***^ | [-0.162,-0.144] | -0.085^***^ | [-0.108,-0.063] |
| Age | 0.016^***^ | [0.016,0.017] | 0.012^***^ | [0.012,0.013] | 0.012^***^ | [0.011,0.013] |
| Ethnicity | 0.005 | [-0.000,0.011] | -0.001 | [-0.007,0.006] | -0.003 | [-0.010,0.004] |
| Social Deprivation | 0.020^***^ | [0.019,0.022] | 0.005^***^ | [0.003,0.006] | 0.005^***^ | [0.003,0.007] |
| BMI |  |  | 0.104^***^ | [0.103,0.105] | 0.100^***^ | [0.097,0.102] |
| Physical Activity |  |  | -0.000^***^ | [-0.000,-0.000] | -0.000^***^ | [-0.000,-0.000] |
| Sedentary Behaviour |  |  | 0.014^***^ | [0.012,0.016] | 0.014^***^ | [0.012,0.016] |
| Sleep Duration |  |  | 0.017^***^ | [0.012,0.022] | 0.016^***^ | [0.011,0.021] |
| Chronotype |  |  | 0.015^***^ | [0.010,0.020] | 0.016^***^ | [0.011,0.021] |
| Shiftwork |  |  | 0.050^***^ | [0.038,0.062] | 0.048^***^ | [0.036,0.061] |
| Smoking |  |  | 0.282^***^ | [0.268,0.297] | 0.281^***^ | [0.267,0.296] |
| Alcohol |  |  | 0.012^***^ | [0.009,0.015] | 0.012^***^ | [0.009,0.016] |
| Vitamin D |  |  |  |  | -0.002^**^ | [-0.004,-0.001] |
| Sex |  |  |  |  | 0.000 | [0.000,0.000] |
| Vitamin D # Sex |  |  |  |  | -0.001^***^ | [-0.002,-0.001] |
| Vitamin D |  |  |  |  | 0.000 | [0.000,0.000] |
| BMI |  |  |  |  | 0.000 | [0.000,0.000] |
| Vitamin D # BMI |  |  |  |  | 0.000^***^ | [0.000,0.000] |
| Outdoor Temperature |  |  |  |  | -0.000 | [-0.002,0.002] |
| Time of Day |  |  |  |  | -0.004^***^ | [-0.005,-0.002] |
| Blood Analyser |  |  |  |  | -0.000 | [-0.004,0.004] |
| Assessment Centre |  |  |  |  | -0.001^**^ | [-0.002,-0.000] |
| Observations | 303707 |  | 172476 |  | 161492 |  |
| *R*^2^ | 0.019 |  | 0.223 |  | 0.222 |  |

95% confidence intervals in brackets

^*^ *p* < 0.05, ^**^ *p* < 0.01, ^***^ *p* < 0.001

| WBC | Model 1 |  | Model 2 |  | Model 3 |  |
| --- | --- | --- | --- | --- | --- | --- |
| Daylength | -0.011^***^ | [-0.013,-0.009] | -0.008^***^ | [-0.011,-0.006] | -0.011^***^ | [-0.015,-0.007] |
| Sex | 0.005 | [-0.007,0.016] | -0.181^***^ | [-0.197,-0.166] | -0.186^***^ | [-0.202,-0.171] |
| Age | 0.002^***^ | [0.002,0.003] | -0.003^***^ | [-0.004,-0.002] | -0.005^***^ | [-0.006,-0.004] |
| Ethnicity | -0.098^***^ | [-0.107,-0.090] | -0.094^***^ | [-0.105,-0.083] | -0.142^***^ | [-0.153,-0.130] |
| Social Deprivation | 0.033^***^ | [0.031,0.035] | 0.006^***^ | [0.004,0.009] | 0.001 | [-0.002,0.003] |
| BMI |  |  | 0.068^***^ | [0.066,0.069] | 0.064^***^ | [0.062,0.066] |
| Physical Activity |  |  | -0.000^*^ | [-0.000,-0.000] | -0.000^**^ | [-0.000,-0.000] |
| Sedentary Behaviour |  |  | 0.027^***^ | [0.023,0.030] | 0.027^***^ | [0.024,0.030] |
| Sleep Duration |  |  | 0.031^***^ | [0.023,0.039] | 0.030^***^ | [0.022,0.038] |
| Chronotype |  |  | -0.013^**^ | [-0.022,-0.005] | -0.033^***^ | [-0.041,-0.025] |
| Shiftwork |  |  | -0.000 | [-0.021,0.020] | 0.027^*^ | [0.006,0.047] |
| Smoking |  |  | 1.223^***^ | [1.198,1.248] | 1.205^***^ | [1.180,1.230] |
| Alcohol |  |  | 0.028^***^ | [0.022,0.033] | 0.025^***^ | [0.019,0.031] |
| Vitamin D |  |  |  |  | -0.003^***^ | [-0.004,-0.003] |
| Outdoor Temperature |  |  |  |  | 0.001 | [-0.001,0.004] |
| Time of Day |  |  |  |  | 0.142^***^ | [0.139,0.144] |
| Blood Analyser |  |  |  |  | -0.016^***^ | [-0.023,-0.009] |
| Assessment Centre |  |  |  |  | 0.012^***^ | [0.011,0.014] |
| Observations | 309516 |  | 175753 |  | 161746 |  |
| *R*^2^ | 0.005 |  | 0.089 |  | 0.163 |  |

95% confidence intervals in brackets

^*^ *p* < 0.05, ^**^ *p* < 0.01, ^***^ *p* < 0.001

Table S7:	Summary descriptive (mean (sd)) statistics for antibody titres by season

|  | **Spring** | **Summer** | **Autumn** | **Winter** | **p-value** |
| --- | --- | --- | --- | --- | --- |
|  | n=1,617 | n=1,757 | n=1,602 | n=1,216 |  |
| **GG** | 3051 (3002) | 3235 (3010) | 3078 (2956) | 3147 (3051) | 0.286 |
| **MGG** | 250 (801) | 307 (965) | 235 (723) | 259 (839) | 0.071 |
| **GEGL** | 916 (1089) | 979 (1185) | 972 (1188) | 965 (1210) | 0.402 |
| **VCAp18** | 7258 (3552) | 7227 (3332) | 7179 (3379) | 7125 (3416) | 0.748 |
| **EBNA_1** | 4294 (3114) | 4364 (3033) | 4513 (3144) | 4410 (3119) | 0.235 |
| **ZEBRA** | 2352 (1989) | 2300 (1897) | 2376 (2013) | 2300 (1916) | 0.619 |
| **EA_D** | 2635 (2687) | 2474 (2508) | 2574 (2550) | 2456 (2583) | 0.179 |
| **PP150** | 1580 (2011) | 1575 (2018) | 1622 (2035) | 1557 (1965) | 0.842 |
| **PP52** | 3094 (3383) | 3036 (3361) | 3276 (3492) | 3095 (3405) | 0.208 |
| **PP28** | 1315 (1672) | 1310 (1657) | 1362 (1686) | 1339 (1639) | 0.796 |
| **IELA** | 293 (325) | 315 (352) | 297 (391) | 317 (370) | 0.160 |
| **IE1B** | 556 (954) | 557 (945) | 486 (800) | 533 (912) | 0.090 |
| **P101K** | 145 (447) | 169 (577) | 167 (477) | 133 (393) | 0.142 |
| **U14** | 868 (838) | 865 (781) | 877 (833) | 850 (775) | 0.849 |
| **LANA** | 79.4 (504) | 82.0 (545) | 107 (737) | 66.8 (415) | 0.282 |
| **K81** | 74.6 (177) | 74.5 (123) | 66.8 (69.9) | 70.7 (104) | 0.248 |
| **HBC** | 66.8 (567) | 71.4 (617) | 57.4 (489) | 71.1 (643) | 0.898 |
| **HBE** | 90.9 (530) | 95.9 (587) | 90.9 (505) | 101 (638) | 0.960 |
| **COREHEPC** | 34.4 (350) | 30.6 (291) | 18.3 (132) | 14.7 (90.8) | 0.094 |
| **NS3** | 54.2 (344) | 50.8 (299) | 40.7 (110) | 38.0 (49.6) | 0.213 |
| **P22** | 73.4 (164) | 75.7 (169) | 69.1 (173) | 73.4 (181) | 0.731 |
| **SAG1** | 124 (135) | 128 (187) | 122 (127) | 125 (157) | 0.747 |
| **HTLV1_GAG** | 280 (348) | 303 (357) | 291 (358) | 288 (351) | 0.301 |
| **HTLV1_ENV** | 32.4 (32.8) | 34.8 (43.1) | 30.6 (26.6) | 32.9 (31.4) | 0.005 |
| **HIV1_GAG** | 117 (273) | 128 (266) | 114 (258) | 116 (248) | 0.464 |
| **HIV1_ENV** | 44.5 (39.4) | 48.1 (56.7) | 43.7 (32.3) | 43.7 (38.3) | 0.008 |
| **BKVP1** | 3781 (2703) | 3744 (2700) | 3807 (2695) | 3799 (2652) | 0.910 |
| **JCVP1** | 754 (1103) | 818 (1205) | 791 (1227) | 762 (1093) | 0.379 |
| **MCVP1** | 2209 (2386) | 2147 (2439) | 2333 (2519) | 2237 (2487) | 0.175 |
| **L1_T16** | 69.1 (185) | 80.5 (247) | 66.0 (172) | 70.5 (161) | 0.162 |
| **E6** | 26.0 (193) | 35.8 (276) | 32.6 (329) | 52.3 (488) | 0.193 |
| **E7** | 36.9 (127) | 46.2 (213) | 33.0 (85.3) | 55.2 (369) | 0.028 |
| **L1_T18** | 54.0 (113) | 58.5 (166) | 57.7 (152) | 56.6 (118) | 0.811 |
| **MOMP_D** | 111 (402) | 145 (469) | 156 (552) | 158 (508) | 0.023 |
| **MOMP_A** | 62.7 (223) | 83.3 (306) | 83.1 (341) | 90.0 (314) | 0.067 |
| **TARP_F1** | 168 (585) | 200 (621) | 194 (651) | 211 (672) | 0.294 |
| **TARP_F2** | 178 (523) | 212 (572) | 198 (558) | 231 (661) | 0.095 |
| **PORB** | 21.2 (43.3) | 24.2 (76.6) | 23.4 (60.1) | 22.9 (39.3) | 0.510 |
| **PGP3** | 509 (1489) | 654 (1678) | 617 (1588) | 673 (1741) | 0.024 |
| **VACA** | 178 (631) | 198 (741) | 191 (727) | 161 (558) | 0.489 |
| **OMP** | 512 (1185) | 526 (1201) | 544 (1274) | 476 (1159) | 0.514 |
| **GROEL** | 1041 (2299) | 1091 (2288) | 1019 (2230) | 962 (2242) | 0.487 |
| **CAT** | 562 (1798) | 619 (1964) | 551 (1775) | 543 (1809) | 0.632 |
| **UREA_AT** | 436 (1438) | 436 (1412) | 440 (1428) | 330 (1113) | 0.114 |
| **CAGA** | 952 (2294) | 1039 (2379) | 898 (2194) | 900 (2136) | 0.535 |

GG	1gG antigen for Herpes Simplex virus-1

MGG	2mgG unique antigen for Herpes Simplex virus-2

GEGL	gE / gI antigen for Varicella Zoster Virus

VCAp18	VCA p18 antigen for Epstein-Barr Virus

EBNA_1	EBNA-1 antigen for Epstein-Barr Virus

ZEBRA	ZEBRA antigen for Epstein-Barr Virus 23005-0.0

EA_D	EA-D antigen for Epstein-Barr Virus 23006-0.0

PP150	pp150 Nter antigen for Human Cytomegalovirus 23007-0.0

PP52	pp 52 antigen for Human Cytomegalovirus 23008-0.0

PP28	pp 28 antigen for Human Cytomegalovirus 23009-0.0

IELA	IE1A antigen for Human Herpesvirus-6 23010-0.0

IE1B	IE1B antigen for Human Herpesvirus-6 23011-0.0

P101K	p101 k antigen for Human Herpesvirus-6 23012-0.0

U14	U14 antigen for Human Herpesvirus-7 23013-0.0

LANA	LANA antigen for Kaposi's Sarcoma-Associated Herpesvirus 23014-0.0

K81	K8.1 antigen for Kaposi's Sarcoma-Associated Herpesvirus 23015-0.0

HBC	HBc antigen for Hepatitis B Virus 23016-0.0

HBE	HBe antigen for Hepatitis B Virus 23017-0.0

COREHEPC	Core antigen for Hepatitis C Virus 23018-0.0

NS3	NS3 antigen for Hepatitis C Virus 23019-0.0

P22	p22 antigen for Toxoplasma gondii 23020-0.0

SAG1	sag1 antigen for Toxoplasma gondii 23021-0.0

HTLV1_GAG	HTLV-1 gag antigen for Human T-Lymphotropic Virus 1 23022-0.0

HTLV1_ENV	HTLV-1 env antigen for Human T-Lymphotropic Virus 1 23023-0.0

HIV1_GAG	HIV-1 gag antigen for Human Immunodeficiency Virus 23024-0.0

HIV1_ENV	HIV-1 env antigen for Human Immunodeficiency Virus 23025-0.0

BKVP1 BK	VP1 antigen for Human Polyomavirus BKV 23026-0.0

JCVP1 JC	VP1 antigen for Human Polyomavirus JCV 23027-0.0

MCVP1 MC	VP1 antigen for Merkel Cell Polyomavirus 23028-0.0

L1_T16	L1 antigen for Human Papillomavirus type-16 23029-0.0

E6	E6 antigen for Human Papillomavirus type-16 23030-0.0

E7	E7 antigen for Human Papillomavirus type-16 23031-0.0

L1_T18	L1 antigen for Human Papillomavirus type-18 23032-0.0

MOMP_D	momp D antigen for Chlamydia trachomatis 23033-0.0

MOMP_A	momp A antigen for Chlamydia trachomatis 23034-0.0

TARP_F1	tarp-D F1 antigen for Chlamydia trachomatis 23035-0.0

TARP_F2	tarp-D F2 antigen for Chlamydia trachomatis 23036-0.0

PORB	PorB antigen for Chlamydia trachomatis 23037-0.0

PGP3	pGP3 antigen for Chlamydia trachomatis 23038-0.0

VACA	VacA antigen for Helicobacter pylori 23040-0.0

OMP	OMP antigen for Helicobacter pylori 23041-0.0

GROEL	GroEL antigen for Helicobacter pylori 23042-0.0

CAT	Catalase antigen for Helicobacter pylori 23043-0.0

UREA_	AT	UreA antigen for Helicobacter pylori 23044-0.0

CAGA	CagA antigen for Helicobacter pylori 23039-0.0

Table S8	Summary descriptive statistics (mean, sd) by time of day for antigen titres.

|  | **Time of Day** | | | | | | | | | | |  |
| --- | --- | --- | --- | --- | --- | --- | --- | --- | --- | --- | --- | --- |
|  | **9** | **10** | **11** | **12** | **13** | **14** | **15** | **16** | **17** | **18** | **19** | **p-value*** |
|  | n=334 | n=512 | n=591 | n=581 | n=661 | n=699 | n=675 | n=655 | n=596 | n=587 | n=246 |  |
| **GG** | 2985 (2985) | 3016 (3032) | 3105 (2879) | 3065 (2941) | 3003 (3001) | 3184 (3031) | 3294 (3040) | 3226 (3115) | 3091 (2895) | 3256 (3058) | 3134 (3063) | 0.722 |
| **MGG** | 294 (930) | 298 (938) | 301 (939) | 285 (902) | 299 (833) | 287 (927) | 243 (757) | 202 (640) | 242 (786) | 243 (815) | 176 (575) | 0.278 |
| **GEGL** | 1025 (1243) | 957 (1178) | 961 (1147) | 949 (1104) | 955 (1158) | 975 (1125) | 1002 (1310) | 928 (1101) | 936 (1191) | 935 (1162) | 924 (1131) | 0.973 |
| **VCAp18** | 6978 (3491) | 7229 (3327) | 7261 (3373) | 7280 (3512) | 7291 (3419) | 7184 (3540) | 7335 (3419) | 7215 (3478) | 7106 (3380) | 6805 (3288) | 7538 (3320) | 0.167 |
| **EBNA_1** | 4230 (3100) | 4493 (3081) | 4420 (2978) | 4309 (3210) | 4535 (3136) | 4221 (3064) | 4493 (2995) | 4378 (3242) | 4467 (3103) | 4140 (3017) | 4567 (3282) | 0.348 |
| **ZEBRA** | 2328 (2066) | 2262 (1863) | 2381 (1934) | 2390 (1904) | 2300 (1954) | 2329 (1946) | 2412 (1966) | 2424 (2088) | 2403 (1987) | 2056 (1878) | 2351 (1858) | 0.079 |
| **EA_D** | 2332 (2438) | 2511 (2597) | 2434 (2491) | 2592 (2515) | 2509 (2585) | 2570 (2693) | 2699 (2612) | 2551 (2613) | 2654 (2607) | 2380 (2540) | 2551 (2672) | 0.474 |
| **PP150** | 1408 (1884) | 1611 (2053) | 1500 (1940) | 1557 (2034) | 1823 (2065) | 1698 (2123) | 1719 (2092) | 1525 (1920) | 1558 (2034) | 1367 (1856) | 1500 (1897) | 0.003 |
| **PP52** | 2646 (3220) | 3082 (3412) | 3057 (3384) | 3052 (3406) | 3504 (3396) | 3252 (3444) | 3317 (3470) | 3176 (3468) | 3060 (3443) | 2835 (3263) | 3122 (3450) | 0.010 |
| **PP28** | 1129 (1465) | 1340 (1706) | 1352 (1734) | 1275 (1602) | 1476 (1669) | 1431 (1771) | 1364 (1619) | 1355 (1662) | 1254 (1562) | 1229 (1689) | 1291 (1703) | 0.069 |
| **IELA** | 286 (272) | 331 (477) | 308 (366) | 299 (358) | 326 (395) | 305 (359) | 281 (327) | 282 (308) | 331 (398) | 292 (282) | 308 (343) | 0.102 |
| **IE1B** | 503 (818) | 566 (991) | 541 (797) | 543 (1035) | 540 (860) | 500 (796) | 516 (892) | 568 (1026) | 505 (841) | 552 (967) | 534 (934) | 0.940 |
| **P101K** | 107 (314) | 203 (575) | 129 (301) | 140 (384) | 154 (459) | 186 (687) | 161 (602) | 126 (351) | 162 (400) | 177 (566) | 110 (263) | 0.036 |
| **U14** | 893 (797) | 914 (884) | 902 (816) | 837 (754) | 788 (695) | 878 (847) | 936 (893) | 857 (826) | 883 (813) | 818 (742) | 797 (784) | 0.034 |
| **LANA** | 35.6 (99.8) | 61.7 (355) | 113 (823) | 51.1 (367) | 80.2 (445) | 102 (614) | 72.4 (502) | 87.9 (667) | 109 (705) | 88.1 (562) | 138 (701) | 0.366 |
| **K81** | 67.5 (70.3) | 78.0 (172) | 81.7 (224) | 71.8 (165) | 68.4 (60.7) | 68.0 (90.9) | 73.0 (150) | 72.1 (80.5) | 68.5 (55.7) | 67.4 (83.9) | 76.2 (96.2) | 0.645 |
| **HBC** | 56.3 (481) | 88.5 (812) | 37.1 (302) | 39.3 (383) | 78.1 (539) | 83.5 (697) | 67.6 (606) | 46.7 (384) | 87.4 (748) | 58.8 (515) | 98.7 (735) | 0.732 |
| **HBE** | 78.9 (448) | 112 (777) | 77.1 (380) | 84.9 (475) | 98.5 (532) | 104 (665) | 81.4 (534) | 85.4 (399) | 111 (680) | 88.9 (527) | 130 (715) | 0.956 |
| **COREHEPC** | 21.8 (84.7) | 14.0 (44.0) | 26.4 (185) | 27.5 (221) | 27.7 (207) | 37.9 (461) | 21.2 (98.0) | 14.5 (56.4) | 27.0 (306) | 18.0 (147) | 15.4 (33.6) | 0.786 |
| **NS3** | 39.0 (40.5) | 38.3 (61.8) | 48.1 (239) | 56.7 (334) | 47.4 (166) | 51.6 (372) | 35.5 (39.0) | 43.6 (92.2) | 51.7 (335) | 37.7 (40.2) | 42.4 (38.1) | 0.830 |
| **P22** | 69.9 (134) | 66.6 (168) | 89.7 (204) | 76.2 (178) | 76.1 (210) | 72.9 (150) | 69.0 (161) | 65.3 (138) | 67.0 (155) | 72.3 (192) | 82.2 (156) | 0.442 |
| **SAG1** | 123 (147) | 118 (122) | 124 (118) | 123 (140) | 132 (165) | 128 (129) | 122 (135) | 120 (135) | 125 (254) | 119 (143) | 146 (167) | 0.521 |
| **HTLV1_GAG** | 319 (357) | 281 (338) | 287 (351) | 288 (348) | 288 (341) | 287 (351) | 263 (340) | 298 (374) | 305 (371) | 317 (361) | 265 (358) | 0.252 |
| **HTLV1_ENV** | 33.5 (28.4) | 30.6 (26.4) | 32.6 (27.7) | 35.1 (56.2) | 32.2 (30.1) | 32.1 (33.2) | 30.4 (25.5) | 33.3 (38.6) | 33.1 (30.5) | 32.1 (28.1) | 38.5 (49.1) | 0.113 |
| **HIV1_GAG** | 145 (335) | 107 (197) | 112 (226) | 122 (248) | 109 (205) | 120 (274) | 147 (388) | 107 (210) | 113 (232) | 123 (271) | 115 (266) | 0.127 |
| **HIV1_ENV** | 43.0 (27.8) | 43.7 (34.9) | 43.9 (29.6) | 44.9 (50.5) | 48.4 (52.2) | 47.6 (46.0) | 41.9 (29.2) | 43.2 (28.8) | 46.8 (40.5) | 45.9 (72.9) | 46.7 (25.3) | 0.157 |
| **BKVP1** | 3956 (2679) | 3988 (2708) | 3691 (2659) | 3782 (2736) | 3723 (2659) | 3753 (2688) | 3577 (2631) | 3801 (2789) | 3831 (2790) | 3824 (2638) | 3731 (2534) | 0.457 |
| **JCVP1** | 884 (1308) | 677 (987) | 799 (1115) | 785 (1214) | 758 (1070) | 792 (1125) | 739 (1111) | 834 (1253) | 802 (1229) | 806 (1245) | 739 (1115) | 0.417 |
| **MCVP1** | 2241 (2401) | 2085 (2444) | 2255 (2498) | 2288 (2530) | 2260 (2481) | 2208 (2481) | 2266 (2462) | 2345 (2456) | 2084 (2360) | 2243 (2452) | 2143 (2336) | 0.786 |
| **L1_T16** | 70.1 (102) | 61.4 (104) | 61.0 (85.2) | 61.9 (131) | 81.0 (255) | 74.6 (255) | 73.1 (181) | 77.5 (267) | 77.9 (190) | 75.7 (236) | 69.0 (160) | 0.666 |
| **E6** | 40.7 (339) | 36.6 (433) | 27.3 (203) | 19.7 (54.4) | 44.6 (474) | 46.4 (444) | 22.4 (90.6) | 27.0 (188) | 34.8 (222) | 52.0 (419) | 21.7 (50.7) | 0.725 |
| **E7** | 37.1 (81.0) | 30.7 (58.9) | 46.1 (191) | 32.5 (84.3) | 36.4 (105) | 66.1 (456) | 33.2 (95.9) | 41.6 (117) | 44.9 (178) | 40.7 (172) | 29.6 (31.7) | 0.072 |
| **L1_T18** | 53.4 (98.5) | 55.4 (121) | 55.4 (98.0) | 52.8 (149) | 52.7 (110) | 58.1 (116) | 60.7 (211) | 59.4 (175) | 59.8 (132) | 56.3 (141) | 58.1 (112) | 0.992 |
| **MOMP_D** | 171 (493) | 142 (443) | 147 (542) | 141 (477) | 150 (557) | 147 (569) | 132 (445) | 125 (383) | 145 (435) | 136 (486) | 124 (412) | 0.981 |
| **MOMP_A** | 83.7 (242) | 66.5 (229) | 79.1 (256) | 84.3 (280) | 88.5 (403) | 81.4 (387) | 73.5 (282) | 64.8 (176) | 85.2 (249) | 96.7 (395) | 57.5 (144) | 0.714 |
| **TARP_F1** | 185 (520) | 197 (601) | 214 (703) | 201 (638) | 175 (586) | 178 (588) | 202 (693) | 175 (606) | 208 (628) | 211 (708) | 123 (469) | 0.796 |
| **TARP_F2** | 219 (527) | 179 (444) | 206 (573) | 205 (597) | 195 (548) | 191 (546) | 225 (764) | 212 (583) | 192 (505) | 218 (583) | 164 (458) | 0.922 |
| **PORB** | 19.5 (21.6) | 19.4 (31.6) | 27.0 (82.2) | 22.1 (42.0) | 23.6 (46.5) | 21.1 (32.2) | 24.2 (56.7) | 22.0 (44.0) | 23.7 (55.2) | 26.2 (113) | 19.5 (31.2) | 0.431 |
| **PGP3** | 763 (1863) | 579 (1445) | 597 (1461) | 570 (1526) | 603 (1613) | 555 (1582) | 597 (1738) | 580 (1572) | 768 (1842) | 602 (1551) | 470 (1459) | 0.264 |
| **VACA** | 174 (726) | 170 (525) | 149 (580) | 175 (638) | 193 (679) | 181 (672) | 161 (575) | 190 (687) | 179 (680) | 263 (987) | 186 (493) | 0.368 |
| **OMP** | 480 (1189) | 562 (1317) | 421 (997) | 597 (1315) | 533 (1168) | 536 (1287) | 485 (1076) | 514 (1211) | 574 (1405) | 479 (1168) | 410 (869) | 0.288 |
| **GROEL** | 953 (2249) | 936 (2236) | 902 (2114) | 1049 (2271) | 1103 (2254) | 1066 (2316) | 1050 (2250) | 1029 (2286) | 1157 (2389) | 1054 (2285) | 982 (2220) | 0.804 |
| **CAT** | 552 (1889) | 425 (1564) | 599 (1884) | 583 (1836) | 681 (2020) | 636 (1971) | 511 (1684) | 524 (1757) | 675 (2069) | 602 (1851) | 272 (1011) | 0.087 |
| **UREA_AT** | 281 (917) | 391 (1355) | 347 (1190) | 440 (1451) | 496 (1505) | 465 (1569) | 392 (1283) | 356 (1280) | 444 (1322) | 447 (1506) | 499 (1392) | 0.351 |
| **CAGA** | 653 (1943) | 946 (2292) | 832 (2116) | 994 (2349) | 933 (2189) | 946 (2193) | 1107 (2451) | 994 (2266) | 1028 (2327) | 933 (2249) | 1171 (2658) | 0.738 |

GG	1gG antigen for Herpes Simplex virus-1

MGG	2mgG unique antigen for Herpes Simplex virus-2

GEGL	gE / gI antigen for Varicella Zoster Virus

VCAp18	VCA p18 antigen for Epstein-Barr Virus

EBNA_1	EBNA-1 antigen for Epstein-Barr Virus

ZEBRA	ZEBRA antigen for Epstein-Barr Virus 23005-0.0

EA_D	EA-D antigen for Epstein-Barr Virus 23006-0.0

PP150	pp150 Nter antigen for Human Cytomegalovirus 23007-0.0

PP52	pp 52 antigen for Human Cytomegalovirus 23008-0.0

PP28	pp 28 antigen for Human Cytomegalovirus 23009-0.0

IELA	IE1A antigen for Human Herpesvirus-6 23010-0.0

IE1B	IE1B antigen for Human Herpesvirus-6 23011-0.0

P101K	p101 k antigen for Human Herpesvirus-6 23012-0.0

U14	U14 antigen for Human Herpesvirus-7 23013-0.0

LANA	LANA antigen for Kaposi's Sarcoma-Associated Herpesvirus 23014-0.0

K81	K8.1 antigen for Kaposi's Sarcoma-Associated Herpesvirus 23015-0.0

HBC	HBc antigen for Hepatitis B Virus 23016-0.0

HBE	HBe antigen for Hepatitis B Virus 23017-0.0

COREHEPC	Core antigen for Hepatitis C Virus 23018-0.0

NS3	NS3 antigen for Hepatitis C Virus 23019-0.0

P22	p22 antigen for Toxoplasma gondii 23020-0.0

SAG1	sag1 antigen for Toxoplasma gondii 23021-0.0

HTLV1_GAG	HTLV-1 gag antigen for Human T-Lymphotropic Virus 1 23022-0.0

HTLV1_ENV	HTLV-1 env antigen for Human T-Lymphotropic Virus 1 23023-0.0

HIV1_GAG	HIV-1 gag antigen for Human Immunodeficiency Virus 23024-0.0

HIV1_ENV	HIV-1 env antigen for Human Immunodeficiency Virus 23025-0.0

BKVP1 BK	VP1 antigen for Human Polyomavirus BKV 23026-0.0

JCVP1 JC	VP1 antigen for Human Polyomavirus JCV 23027-0.0

MCVP1 MC	VP1 antigen for Merkel Cell Polyomavirus 23028-0.0

L1_T16	L1 antigen for Human Papillomavirus type-16 23029-0.0

E6	E6 antigen for Human Papillomavirus type-16 23030-0.0

E7	E7 antigen for Human Papillomavirus type-16 23031-0.0

L1_T18	L1 antigen for Human Papillomavirus type-18 23032-0.0

MOMP_D	momp D antigen for Chlamydia trachomatis 23033-0.0

MOMP_A	momp A antigen for Chlamydia trachomatis 23034-0.0

TARP_F1	tarp-D F1 antigen for Chlamydia trachomatis 23035-0.0

TARP_F2	tarp-D F2 antigen for Chlamydia trachomatis 23036-0.0

PORB	PorB antigen for Chlamydia trachomatis 23037-0.0

PGP3	pGP3 antigen for Chlamydia trachomatis 23038-0.0

VACA	VacA antigen for Helicobacter pylori 23040-0.0

OMP	OMP antigen for Helicobacter pylori 23041-0.0

GROEL	GroEL antigen for Helicobacter pylori 23042-0.0

CAT	Catalase antigen for Helicobacter pylori 23043-0.0

UREA_	AT	UreA antigen for Helicobacter pylori 23044-0.0

CAGA	CagA antigen for Helicobacter pylori 23039-0.0
